# Cost saving in primary versus tertiary level of reproductive health services in Sana’a, Yemen

**DOI:** 10.1101/2021.08.25.21262373

**Authors:** Abdulkareem Ali Hussein Nassar, Yahia Ahmed Raja’a, Najia Saleh Bahubaishi

## Abstract

**Introduction:** In Yemen, women tend to bypass the nearby primary health level facilities (PHLF) for using vaginal delivery (VD), antenatal care (ANC) and intrauterine device (IUD) services. This study aimed to estimate the cost saving for utilization of VD, ANC and IUD services at PHLF instead of tertiary health level facilities (THLF) in Sana’a.

**Methods:** A comparative cross sectional study design was conducted in 2013. It was used to estimate the costs from the patient’s perspective. A structured questionnaire was used to collect data. Descriptive analyses were performed. P value <0.05 was considered as the cut point for significance. The SPSS version 17 was used.

**Results:** A total of 180 women were found. The median of DMC of VD, ANC and IUD services were US$43.9, US$14.8 and US$9.1 at THLF compared with US$19.5, US$0.9 and US$11.2 at PHLF, respectively. The DMC difference of VD, ANC and IUD services between THLF and PHLF was US$24.4, US$13.9 and US$-2.1, respectively. Regarding the DNMC, the median of VD, ANC and IUD services were US$43.1, US$19.1 and US$17.3 at THLF compared with US$14.0, US$0.0 and US$0.0 at PHLF, respectively. The DNMC difference of VD, ANC and IUD service between THLF and PHLF was US$29.1, US$19.1 and US$17.3, respectively. Moreover, the median of INDC for VD, ANC and IUD services were US$23.9, US$9.5 and US$10.4 at THLF compared with US$7.9, US$1.6 and US$1.1 at PHLF, respectively. The INDC difference of VD, ANC and IUD service between THLF and PHLF was US$16.0, US$7.9 and US$9.3.

**Conclusion:** The study found the utilization of VD, ANC and IUD services at PHLF instead of THLF is a considerable cost saving for families. Therefore, shifting the utilization of services from THLF to PHLF reduces the financial burden affecting individuals, families and their productivity. Implementation of the health referral system is recommended.

**Key questions:** *What is already known?:*

➢ Women tend remarkably to bypass the nearby primary reproductive health services that are rising the expenses and increasing the burden borne by individuals, families and their productivity.
➢ Few studies have estimated the cost saving for utilization of primary instead tertiary level of reproductive health services in the world, while no such study was conducted in Yemen.

*What are the new findings?:*

➢ A significant difference in the expenses on reproductive health services between the primary and tertiary health level facilities.
➢ A significant cost is saved for women and families as a result of utilization of primary instead tertiary level of reproductive health services.

*What do the new findings imply?:*

➢ The primary health level facilities reduce the financial burden affecting individuals, families and their productivity.
➢ This study provides evidence for decision-makers to implement the health referral system and protect the families from incurring the high expenditure.

## Introduction

Global initiatives have embraced ambitious new goals to reduce maternal mortality through improving primary health care and achieving health coverage. However, maternal mortality remains the second leading cause of death among women of reproductive age. In 2017, an estimated 295,000 maternal deaths, almost all of them in the developing countries[1, 2].

Despite the global efforts to improve primary health care services, women still tend to bypass the nearby primary reproductive health services (RHS) and it grows frequently in developing countries[3, 4]. Previous studies showed that bypassing the primary health level facilities (PHLF) to use the antenatal care (ANC)[3, 5, 6, 7, 8, 9], vaginal delivery (VD)[3, 5, 6, 7, 9, 10, 11, 12, 13, 14, 15, 16, 17], and family planning services[18] at higher levels even if the care available at the nearby PHLF. These studies indicated that the direct medical cost (DMC), direct non-medical cost (DNMC), and indirect costs (INDC) of RHS are different between PHLF and tertiary health level facilities (THLF). The bypassing transfers the health care expenditures away from DMC to DNMC and INDC. The women need more accompanying individuals, long distance and absence from their work to get RHS at higher levels, which is reflected on borne more expenses such as transportation, food, lost wages[3, 5, 6, 8, 9, 10, 11, 14, 15, 16, 17,18]. Therefore, this phenomenon increases the financial burden on families, underutilizes the nearby PHLF and overloads the higher level facilities. Conversely, reducing the bypassing would reduce the financial burden borne by families associated with traveling further among bypassers[4].

Yemen National Health and Demographic Survey (YNHDS) in 2013 indicated the maternal health indicators have improved in recent years, however it is still low. The maternal mortality ratio is 148 deaths per 100,000 live births, 60% of Yemeni women received ANC, 45% of deliveries performed by skilled health workers and 29% married women are currently using a modern method of family planning[19]. In 2011, Yemen aimed in the national reproductive health policy to improve access to high quality reproductive health services as well. The number of health facilities has significantly increased in all Yemeni districts, nearly 80% of the health facilities are in the rural areas[20]. However, women tend remarkably to go to hospitals seeking health care that is growing demand on health services, rising the expenses on health care, overloading of hospitals and increasing the burden of expenses on women that can be reduced by utilizing RHS at PHLF in their regions. Moreover, the burden of household spending on healthcare has escalated over the past 10 years due to political and military conflicts. The conflict started as demonstrations in 2011, followed by internal fighting. The intervention of the Saudi-led coalition in 2015 and the blockade imposed so far have worsened the households’ economic situation, such as stopping salaries and increasing the prices of commodities and fuel[1]. Consequently, understanding of the economic aspect and quality of RHS is critical especially during collapsing economic situation with limited resources and growing costs. The community needs to be aware of the size of healthcare expenses in different health levels and cost saving due to avoid bypassing the nearby PHLF, particularly in rural areas where nearly 70% of the total population live[1]. Estimating the cost of RHS can provide valuable information for decision-makers about the size of problems in the health system, assess the resources used in facilities, and suggest improving the efficiency of health services.

In Yemen no such study has been conducted to estimate the cost of VD, ANC and family planning only intrauterine device (IUD) services. This study aimed to estimate the DMC, DNMC and INDC saving for utilization of VD, ANC and IUD services at PHLF instead of THLF in Sana’a city and its surrounding districts, and identify the reasons for not using the PHLF.

## Methods

### Study design and setting

A comparative cross sectional study design was conducted among RH clients/patients who live in Sana’a governorate in 2013. It was used to estimate the costs of RHS at THLF compared with PHLF in Sana’a city and its surrounding districts (Sana’a governorate). The cost was estimated from the patient’s perspective (out-of-pocket expenses born by RH clients/patients and accompanying individuals). The PHLF including health centers provide primary health care services, which are preventive, diagnostic and therapeutic health services, and represents the link between the health units and district Hospitals. The RHS is one of these provided services in PHLF, such as VD, ANC and IUD services. The THLF provides more complex and specialized services to patients who are referred from lower levels of the health system where such services are not available. However, the hospitals in Yemen provide primary, secondary, and tertiary health care services, in contrast to its function.

### Selected health facilities and services

A total of 8 health centers in Sana’a governorate and 3 hospitals in Sana’a city were selected. Hezam, Walan, Bani Mansor, Al Rekh, Bait Ghofr, Al Kebs, Al Aghmoor and Ghaiman Health Centers were selected from Sana’a governorate. Availability of the selected RH services and clients (Sana’a governorate’s PHP office, Reproductive health department. Reproductive health services report. 2010) and the geographically surrounding districts that aren’t far from Sana’a city and closer than any other city (Ministry of Public Health and Population, Researches and data administration, Map of health centers in Sana’a governorate, 2010) were taken into consideration when selecting these health centers. While Al-Thawra, Al-Sab’een and Al-Kuwait Hospitals were selected. These three hospitals are considered the major public hospitals in Sana’a city. Three RHS (VD, ANC and IUD) were involved in this study. The services were selected according to the following considerations: the services are more available and common in PHLF (Sana’a governorate’s Public Health and Population office, Reproductive health department. Reproductive health services report. 2010) IUD is the second of family planning methods used and preferred by most women especially in rural areas (25.6%)[19]. As well the services are more cost and measurable compared to other RHS at PHLF.

### Sample Size

A sample size of 180 RH clients/patients were enrolled. It was classified equally according to the RHS into 60 women who came for VD, 60 for ANC and 60 for IUD. They were allocated equally into PHLF and THLF groups.

As a result of political conflicts and the limited financial resources, a convenience sample size was determined as 180 from PHLF and THLF groups, and for the selected three RHS. A sample size of 30 is considered as a minimum statistical number and sufficient to estimate the quantitative variables (n ≥ 30 is the guideline for quantitative variables)[21].

### Inclusion and exclusion criteria

This study included women aged (15 – 45 years), who lived in the surrounding districts of Sana’a city and came for VD, ANC or IUD. The accompanying adult individuals (male and female) were included. As well those who agree to participate in this study. While women who were subjected to cesarean section, referred from PHLF to THLF and came for family planning (pills, condoms, injection, tubal ligation and implants) were excluded. women in the post-menopausal state or who lived in districts other than the surrounding districts of Sana’a city were also excluded. As well as accompanying children and those who disagree to participate in this study.

### Data collection and measurements

A structured questionnaire was quoted from literatures[5, 11, 12] and then reconstructed in the English language according to the objectives of the study. It was translated into the Arabic language. The data was collected through a face-to-face interview. The questionnaire was tested and reviewed by two experts, to ensure simplicity and clarity of questions. It consisted of questions to collect data related to socio-demographic characteristics, DMC and DNMC, and INDC. The socio-demographic characteristics include age, education and working status, and residence of RH clients/patients and accompanying individuals (is one or more persons who associated clients to health facilities). The DMC was elicited through questions related to the cost of clinical visits, diagnostic procedures (laboratory test and ultrasound), therapeutic procedures (delivery and IUD insertion or removal), medication costs (drugs, IUD items, tetanus toxoid vaccine and hepatitis B virus vaccine), and other expenses related to medical care, such as baby milk and diapers costs. The RH clients/patients and accompanying individuals were asked questions on the following DNMC; transportation (the trips from home to the health facility and back), accommodation, eating, khat and other costs like tips, clothes for baby or mother, blanket, cigarette and phone call. To estimate the INDC (lost wages due to absence from work), RH clients/patients and accompanying individuals were asked questions related to the time lost (time spent from leaving until return to home), and working status and wage. One more open-ended question for the THLF group to elicit data about the reasons for not using the nearby or adjacent PHLF.

### Data processing and analysis

Data were entered and cleaned in an Excel program and analyzed by the Statistical Package for the Social Sciences (SPSS) version 17. All costs are presented in the local currency, Yemeni Riyal (YER), and change to US$ using the average exchange rate for 2013 (1 US$ = YER 214.89). The wage rate of housewives was estimated according to the minimum limit of wage in Yemen, 20,000 YER per month (Ministry of Civil Service and Pensions, The law of Jobs, Wages and Salaries System. Law No. (43). Article (38). Paragraph (E). 2005. p. 10). The wages of those who work without earnings derived from the replacement cost method which uses the wages of persons who work to pay for these tasks, as a measure of their value. The INDC was calculated by multiplying the wage per minute by the time lost in minutes. Cost saving is calculated as the difference between the cost in PHLF and THLF. The responses to open-ended questions were grouped, classified and summarized into seven themes. Descriptive analyses were performed to calculate the percentage, median, range and interquartile range (IQR) as quartile 1 and quartile 3. As well as the median difference was calculated to compare the costs between PHLF and THLF groups. The mean and SD were used as an additional measure. The Mann-Whitney U test was used to compare the differences between PHLF and THLF groups. P value < 0.05 was considered statistically significant.

## Results

A total of 180 RH clients/patients were enrolled from PHLF and THLF, equally. Sixty women were attended for each RHS (VD, ANC and IUD services). In addition, 246 accompanying individuals attended with RH clients/patients were interviewed to estimate the DNMC and INDC.

### Socio-demographic characteristics

Table 1 shows the socio-demographic characteristics of the RH clients/patients. The median (IQR) age of VD clients/patients among PHLF and THLF groups were 25 (22 and 30) years and 26 (20 and 30) years, respectively (p value = 0.800). The percentage of illiterate was 90% among PHLF and 80% THLF groups. All the VD clients were housewives in the PHLF group and 93% were housewives in the THLF group.

**Table 1:**
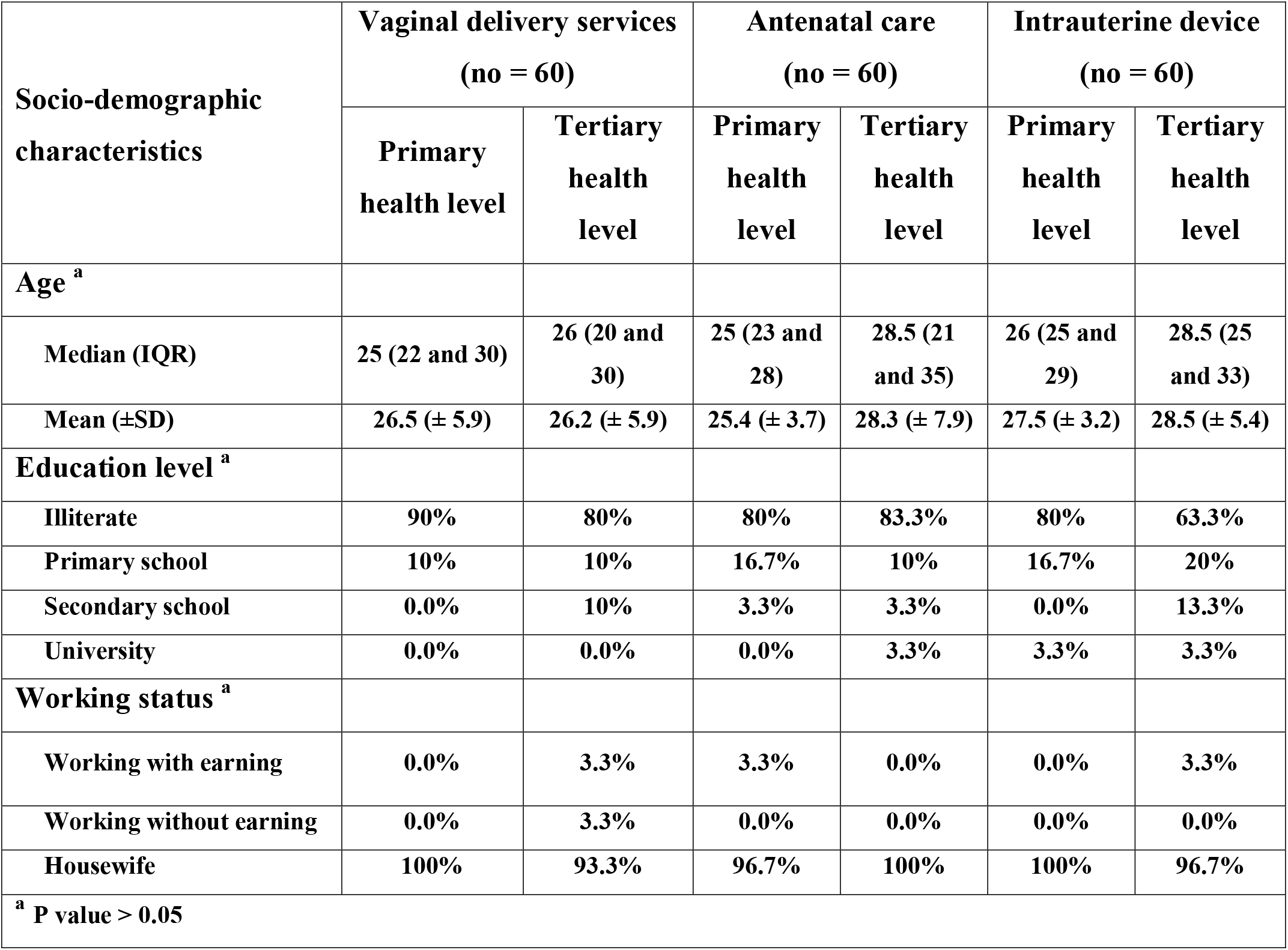
Socio-demographic characteristics of reproductive health clients/patients in primary and tertiary health levels.

| Socio-demographic characteristics | Vaginal delivery services<br>(no = 60) |  | Antenatal care<br>(no = 60) |  | Intrauterine device<br>(no = 60) |  |
| --- | --- | --- | --- | --- | --- | --- |
|  | Primary health level | Tertiary health level | Primary health level | Tertiary health level | Primary health level | Tertiary health level |
| Age <sup>a</sup> |  |  |  |  |  |  |
| Median (IQR) | 25 (22 and 30) | 26 (20 and 30) | 25 (23 and 28) | 28.5 (21 and 35) | 26 (25 and 29) | 28.5 (25 and 33) |
| Mean (±SD) | 26.5 (± 5.9) | 26.2 (± 5.9) | 25.4 (± 3.7) | 28.3 (± 7.9) | 27.5 (± 3.2) | 28.5 (± 5.4) |
| Education level <sup>a</sup> |  |  |  |  |  |  |
| Illiterate | 90% | 80% | 80% | 83.3% | 80% | 63.3% |
| Primary school | 10% | 10% | 16.7% | 10% | 16.7% | 20% |
| Secondary school | 0.0% | 10% | 3.3% | 3.3% | 0.0% | 13.3% |
| University | 0.0% | 0.0% | 0.0% | 3.3% | 3.3% | 3.3% |
| Working status <sup>a</sup> |  |  |  |  |  |  |
| Working with earning | 0.0% | 3.3% | 3.3% | 0.0% | 0.0% | 3.3% |
| Working without earning | 0.0% | 3.3% | 0.0% | 0.0% | 0.0% | 0.0% |
| Housewife | 100% | 93.3% | 96.7% | 100% | 100% | 96.7% |
| <sup>a</sup> P value > 0.05 |  |  |  |  |  |  |

For ANC, the median (IQR) of age among PHLF and THLF groups were 25 (23 and 28) years and 28.5 (21 and 35) years, respectively (p value = 0.186). The percentage of illiterate were 80% and 83% among PHLF and THLF groups, respectively. almost 97% were housewives in the PHLF group and 100% were housewives in the THLF group.

Moreover, the median (IQR) age of IUD clients among PHLF and THLF groups were 26 (25 and 29) years and 28.5 (25 and 33), respectively (p value = 0.467). The percentage of illiterate were 80% and 63% among PHLF and THLF groups, respectively.

All IUD clients were housewives in the PHLF group and 97% were housewives in the THLF group.

Table 2 shows the distribution of the costs of RHS in PHLF and THLF. The overall costs median (IQR) of VD, ANC and IUD services were US$ 114.1 (91.6 and 140.5), US$ 43.4 (33.6 and 67.8), and US$ 37.7 (31.9 and 52.6) at THLF compared with US$ 42.8 (26.6 and 53.9), US$ 3.1 (1.6 and 6.1) and US$ 14.2(11.1 and 18.4) at PHLF, respectively. The overall cost difference between THLF and PHLF groups was US$ 71.3 (166%) for VD, US$ 40.3 (1,300%) for ANC and US$ 23.5 (165%) for IUD services, respectively (with p value < 0.0001).

**Table 2:**
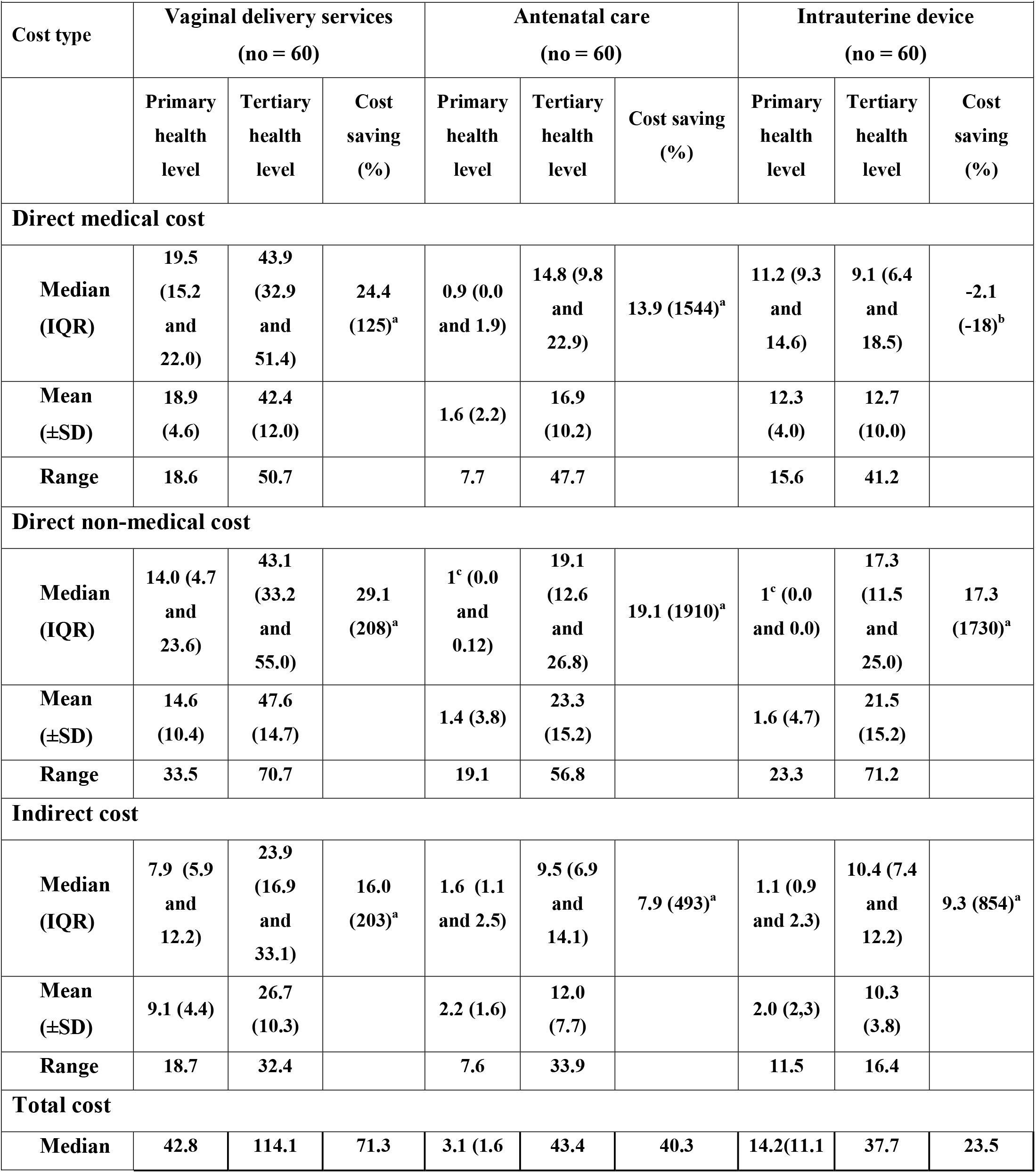
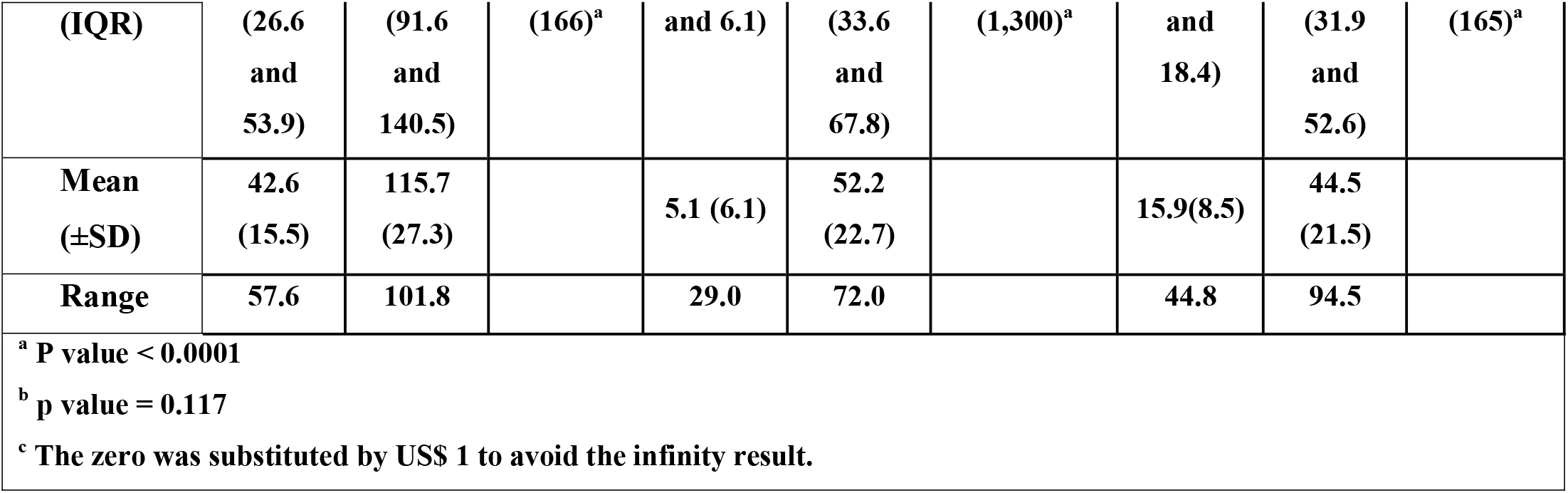
Distribution the costs of reproductive health services in primary and tertiary health level facilities.

### Direct medical cost

Table 2 shows the median (IQR) of DMC for VD service was US$ 43.9 (32.9 and 51.4) at THLF compared with US$ 19.5 (15.2 and 22.0) at PHLF. Similarly, the median (IQR) of DMC for ANC service in THLF and PHLF were US$ 14.8 (9.8 and 22.9) and US$ 0.9 (0.0 and 1.9), respectively. However, the median (IQR) of DMC for IUD service was US$ 9.1 (6.4 and 18.5) in THLF and US$ 11.2 (9.3 and 14.6) in PHLF. The DMC difference between THLF and PHLF groups was US$ 24.4 (p value < 0.0001) for VD service, US$ 13.9 (p value < 0.0001) for ANC service and US$ -2.1 (p value = 0.117) for IUD service.

### Direct non-medical cost

Regarding the DNMC, the median (IQR) of DNMC of VD was US$ 43.1 (33.2 and 55.0) in THLF compared with US$ 14.0 (4.7 and 23.6) in PHLF. For ANC service, the median (IQR) was US$ 19.1 (12.6 and 26.8) at THLF compared with US$ 0.0 (0.0 and 0.12) at PHLF. Moreover, the median (IQR) of DNMC for IUD service were US$ 17.3 (11.5 and 25.0) in THLF compared with US$ 0.0 (0.0 and 0.0) in PHLF. Therefore, The DNMC difference between THLF and PHLF groups was US$ 29.1 (p value < 0.0001) for VD service, US$ 19.1 (p value < 0.0001) for ANC service and US$ 17.3 (p value < 0.0001) for IUD service (table 2).

### Indirect cost

The median (IQR) of INDC for VD, ANC and IUD services were US$ 23.9 (16.9 and 33.1), US$ 9.5 (6.9 and 14.1), and US$ 10.4 (7.4 and 12.2) at THLF compared with US$ 7.9 (5.9 and 12.2), US$ 1.6 (1.1 and 2.5) and US$ 1.1 (0.9 and 2.3) at PHLF, respectively. Therefore, the INDC difference between THLF and PHLF groups was US$ 16.0 (p value < 0.0001) for VD service, US$ 7.9 (p value < 0.0001) ANC service and US$ 9.3 (p value < 0.0001) for IUD service (table 2).

### Reasons for not using primary health level facilities

Table 3 shows the reasons for not using RHS at PHLF. The reasons for not using VD services were unavailability of gynecologists at PHLF (28%), followed by carelessness (19%), and loss of confidence at PHLF (19%). While unskilled health workers (24%), carelessness (18%), unavailability of gynecologists (15%), unavailability of technical resources (15%), and loss of confidence (11%) were the reasons for not using ANC at PHLF. Moreover, the appropriate or free price of IUD services at THLF was the main reason (22%), followed by unskilled health workers (18%) and unavailability of technical resources at PHLF (18%).

**Table 3:** The reasons for not using reproductive health services in primary health level facilities.

|  |  |  |  |  |
| --- | --- | --- | --- | --- |
| **Reasons** | **Vaginal delivery**<br>**services**<br>**No. (%)** | **Antenatal**<br>**care**<br>**No. (%)** | **Intrauterine**<br>**device**<br>**No. (%)** | **Total**<br>**No. (%)** |
| **Unavailability of gynecologists** | **12 (28)** | **5 (15)** | **4 (12)** | **21 (19)** |
| **Carelessness in health centers** | **8 (19)** | **6 (18)** | **2 (6)** | **16 (15)** |
| **Loss of confidence** | **8 (19)** | **4 (11)** | **5 (15)** | **17 (15.5)** |
| **Unavailability of technical resources** | **6 (14)** | **5 (15)** | **6 (18)** | **17 (15.5)** |
| **Unskilled health workers** | **5 (11)** | **8 (24)** | **6 (18)** | **19 (17)** |
| **Unavailability\*** | **4 (9)** | **4 (11)** | **3 (9)** | **11 (10)** |
| **Appropriate or free price in hospitals** | **0 (0)** | **2 (6)** | **7 (22)** | **9 (8)** |
| **Total answers** | **43 (100)** | **34 (100)** | **33 (100)** | **110 (100)** |
| **\* Unavailability: include health center is closed or non-existent, unavailability of drugs and the doctors are transferred to other health facilities** |  |  |  |  |

## Discussion

This is the first study on cost saving in primary versus tertiary RHS conducted in Yemen. It revealed RHS costs were significantly different in PHLF and THLF, except for the DMC of IUD service. Therefore, the utilization of PHLF would be a significant saving for RH clients and their families.

Our results indicate that the ages of RH clients at both PHLF and THLF were comparable. As well as Higher illiteracy and unemployment among married women. This is because all the clients come from the same districts of Sana’a governorate who share the same cultural trends of reproductive behavior, education and employment. Our findings revealed that the DMC of VD services was significantly higher at THLF compared to PHLF (p value < 0.0001). The utilization of PHLF instead of THLF saves the family US$ 24.4 (125%) of DMC for VD services. This result is consistent with previous studies in Tanzania[10, 12, 16], Bangladesh[5, 14], India[9, 17], Ghana[3], Malawi[6], Ethiopia[13], three African countries (Burkina Faso, Kenya and Tanzania)[11], Zambia[15], Burkina Faso[22], Vietnam[23], Pakistan[24], Afghanistan[25], and the Democratic Republic of the Congo[26] reported that the DMC of VD was higher at a high level or among bypassers compared to low level or non-bypassers. As well as two systematic reviews studies in low-income and middle-income[2, 7].

Moreover, the study found the DMC of ANC was more than fifteen times higher in THFL compared to PHFL (p value < 0.0001). The DMC saving of one ANC visit at PHLF instead of THLF was US$ 13.9 (1,544%). Our finding similar to the finding of studies in Bangladesh[5, 8, 14], Malawi[6], India[9], Ghana[3], Vietnam[23], and Niger[27] showed that the DMC of ANC was higher at a high level compared to a low level. As well as two systematic reviews studies in low-income and middle-income[2, 7]. In contrast, this study indicates that there is no significant difference between the DMC of IUD at THFL and PHFL (p value = 0.117). This might be because the hospitals don’t usually charge for IUD insertion or removal in contrast to the health centers. This result disagrees with the result of the study in Ghana that reported the family planning services at the THLF were high in cost compared to PHLF[18].

The DMC difference of VD and ANC between PHLF and THLF could be attributed to several possible explanations; the provision of services is a shortage, the laboratory tests are often not requested and the ultrasound is often not available in PHLF as compared to that in THLF. The drug prescription pattern of the specialists (higher professional qualifications) in THLF differs from the less qualified personnel (midwives) in PHLF. As well as the variation in availability and price of drugs.

Regarding the DNMC, this study revealed that the expenses for VD were three times higher in THLF compared to PHLF (p value < 0.0001). Therefore, the DNMC saving as a result of the utilization of the VD services at PHLF rather than THLF is US$ 29.1 (208%). This result agrees with the result of previous studies in Bangladesh[5], Malawi[6], Ghana[3], and Tanzania[12] reported that the DNMC of VD was higher at a high level or among bypassers compared to a low level or non-bypassers. Other studies in Tanzania[10, 16], Zambia[15], and India[9, 17] indicated that the transport costs are higher at a high level or among bypassers.

In addition, the study showed that the utilization of PHLF instead of THLF might save the whole DNMC of ANC and IUD services (p value < 0.0001). Two previous studies in Bangladesh[5, 8, 14], Ghana[3], and Malawi[6] indicated that the DNMC of ANC was higher at a high level or among bypassers compared to a low level or non-bypassers. A study in Ghana reported that the transportation cost for family planning services at the THLF was higher in cost compared to PHLF[18].

The DNMC difference of VD, ANC and IUD services between PHLF and THLF is possibly owing to discrepancies in transportation, food, water and number of accompanying individuals. The far distance of the THLF from the client’s homes pushes them to hire vehicles which costs considerably while most RH clients usually cover the distance to PHLF walking, especially for ANC and IUD services. The RH clients usually have more accompanying persons in THLF than in PHLF and this increases the DNMC in terms of transportation and food, while most RH clients either go alone or with minimal accompanying individuals for PHLF especially in ANC and IUD services. Our findings agree with a previous study in Nepal[28] that showed the expenses on feeding and accommodation are higher among those bypassing PHC facilities.

Furthermore, the result of this study revealed that the INDC for the VD client and her accompanying individuals was three times higher at THLF compared to that at PHLF (p value < 0.0001). The INDC saving as a result of the utilization of the VD services at PHLF rather than THLF is US$ 16.0 (203%). This result is consistent with the results of studies in Bangladesh[5] and Malawi[6], reported that the costs of traveling and waiting time of VD were higher at a high level compared to a low level. A study in Tanzania indicated that bypassers incur a substantial opportunity cost due to long time away from their farming work[10].

Our result found the INDC for the ANC client and her accompanying individuals was six times costlier at THLF compared to that at PHLF (p value < 0.0001). The INDC saving due to utilization of one ANC visit at PHLF was US$ 7.9 (498%). The INDC of ANC at THLF might cover the total expenses of ANC at PHLF. This result agrees with the result of studies Bangladesh[8, 14] reported that the lost wage for ANC visit was greater at a high level compared to a low level. Other study in Malawi[6] indicated that the costs of traveling and waiting time were higher at a high level compared to a low level.

Similarly, The INDC for the IUD client and her accompanying individuals are nine times costlier at THLF compared with PHLF (p value < 0.0001). The PHLF might save US$ 904 (854%) of INDC for the IUD client and her accompanying individuals. A study in Ghana reported that the time value lost for family planning services was higher at a high level compared to a low level[18].

The difference in lost wage for RH clients and their accompanying individuals between PHLF and THLF is possibly due to the discrepancy in the location of these services from the client’s homes and the number of accompanying individuals

Additionally, the qualitative part of this study showed the utilization of RHS at THLF instead of PHLF. The majority of women did not use the VD services at PHLF because of the unavailability of gynecologists, loss of confidence and carelessness in health centers. Four studies in Tanzania reported that the good provider performance or practice[10, 12, 23, 29], a greater trust in health workers[10, 12], and availability of drugs and medical equipment[16, 29] were the main reasons for selecting the facility, while the decreasing cost was little influence[29]. Other study in Vietnam reported that women often trust the professional qualifications of physicians and medical equipment at upper level facilities to give birth[23]. As well as study in Nepal reported that lack of necessary equipment and drugs, lack of skilled health workers and low confidence were the reasons for bypassing[4].

This study found that unskilled health workers were the main reason for not using ANC at PHLF, followed by the carelessness in health centers. However, the appropriate or free price of IUD was the most important reason for selecting THLF. Unskilled health workers and unavailability of technical resources in PHLF were other reasons that make women bypass the PHLF. Although FP methods are provided for free in all PHLF in Yemen, still the patients have to pay the costs, as well as unskilled nurses[20]. A study in Yemen found that rural women’s use of health centers for VD and ANC were limited by their perceived poor quality of services, as indicated by the lack of critical staff, particularly female doctors, equipment, and essential medicines[1]. As well YNHDS indicated that 63% of women report no female provider is a problem of accessing health care[19].

The strength of this study is that the first study on cost saving in the primary versus the tertiary level of RHS conducted in Yemen. In addition to interviews before discharge, the costs paid after discharge were collected by phone when they arrived home. However, it has some limitations which should be considered. First, the cost might be estimated as the minimum economic burden of cost as a result of the utilization of RHS at THLF because it was conducted from the patient perspective. Second, it used a non-probability sampling technique which limits the generalization of our results. Third, because of different types of vehicles (according to fuel type, fuel consumption and car size) and different ways, and lack of fixed price for distance, the private transport cost was estimated by asking about the cost of trips from home to the health facility and back as a measure of their value.

The study found that there is a significant discrepancy in the RHS expenses between the THLF and PHLF. The utilization of VD, ANC and IUD services at PHLF is a considerable cost saving for families. Therefore, shifting the utilization of services from THLF to PHLF reduces the financial burden affecting individuals, families and their productivity. As well as issues related to the quality of RHS such as availability of gynecologists, skilled health workers, technical resources were the possible reasons for bypassing the PHLF.

Efforts are urgently needed to enhance the function of PHLF and protect the families from incurring the high expenditure. Development and implementation of the health referral system between health facility levels, and improving the quality of the health services in PHLF are highly recommended. Further research to estimate the RHS costs from the provider perspective is also recommended.

## Data Availability

Data is available

## Abbreviations

ANC: Antenatal care
DMC: Direct medical cost
DNMC: Direct non-medical cost
INDC: Indirect costs
IQR: Interquartile rang
IUD: Intrauterine device
PHLF: Primary health level facilities
RH: Reproductive health
RHS: Reproductive health services
THLF: Tertiary health level facilities
VD: Vaginal delivery
YER: Yemeni Riyal
YNHDS: Yemen National Health and Demographic Survey

## Acknowledgements

We would like to thank Dr. Tamadhur Al-Sabahi and Nabelah Al-Fakeeh for their assistance in data collection, and Malak Al Sabahi for proofreading our manuscript.

## Authors’ contributions

AAHN was the principal author involved in the concept, design and implementation of the study, interpretation of data, and prepared the draft manuscript. YAR was the main supervisor who analyzed the data and contributed in the concept, design and review of the study. NSB was co-supervisor who contributed in the interpretation of data and review the study. All authors reviewed and approved the final version of the manuscript.

## Availability of data and materials

All relevant data are presented in this paper; and more information can be provided upon reasonable request from the corresponding author.

## Ethics approval and consent to participate

The study was conducted and submitted for the partial fulfillment of the requirements for the degree of Master. It was approved by the Research and Ethics Committee of the Faculty of Medicine and Health Science, Sana’a University. Official letters to conduct this study were sent to The Public Health and Population Offices and the targeted facilities in Sana’a city and governorate. Methods were performed in accordance with the Declaration of Helsinki.

## Patient and public involvement

Informed consent was taken from all participants. Confidentiality of data was assured and ensured.

## Patient consent for publication

Not applicable.

